# Hereditary haemochromatosis beyond liver cancer: increased risk of prostate cancer during an 11-year follow-up

**DOI:** 10.1101/2022.02.28.22271621

**Authors:** Janice L Atkins, Luke C Pilling, Suzy V Torti, Frank M Torti, George A Kuchel, David Melzer

## Abstract

In European ancestry populations, the iron overload disorder Hereditary Haemochromatosis (HH) is predominantly caused by the *HFE* p.C282Y mutation. Male p.C282Y homozygotes have markedly increased hepatic malignancy risks but risks for other cancers in both male and female homozygotes are unclear. We studied data from 451,143 UK Biobank European ancestry community participants (aged 40-70 years; 54.3% female) followed for a mean 11.6 years via hospital admission and national cancer registry data. We estimated risks of incident bladder, blood (with sub-analyses of leukemia and lymphoma), bone, brain, breast, colorectal, kidney, lung, melanoma, oesophageal, ovarian, pancreatic, prostate and stomach cancers in the 2,890 p.C282Y homozygotes, compared to participants without *HFE* mutations. Male p.C282Y homozygotes (12.1% with baseline diagnosed HH) had increased risks of prostate cancer (6.8% versus 5.4% without mutations, hazard ratio HR=1.32, 95% CI=1.07-1.63, p=0.01,). In lifetable estimates from ages 40-75 years, 14.4% of male p.C282Y homozygotes are projected to develop prostate cancer (versus 10.7% without mutations, excess 3.8%, 95% CI = 1.3-6.8). There was no excess of studied cancers in female p.C282Y homozygotes, or in p.C282Y/p.H63D compound heterozygotes of either sex. In a large community sample of male p.C282Y homozygotes, there was a 32% increase in prostate cancer risk, in addition to the known increased liver cancer risk. Those reporting *HFE* genotype status or monitoring p.C282Y homozygous males may need to ensure clinical monitoring for both liver and prostate cancers.

**Novelty and impact of the work:** Male *HFE* p.C282Y homozygotes, at risk of iron overload disorder hereditary haemochromatosis, have known increased liver cancer risks. We have shown, for the first time, that the male p.C282Y homozygotes also have a 32% increased risk of prostate cancer; with an excess proportion of 3.8% by age 75 compared to those without mutations. Those reporting *HFE* genotype status or monitoring p.C282Y homozygous males need to ensure vigilance for both excess liver and prostate cancers.

## Introduction

Hereditary haemochromatosis (HH) is a common genetic disorder of iron overload, predominantly caused by the *HFE* p.C282Y mutation and to a lesser extent the *HFE* p.H63D mutation^1^. The p.C282Y variant is carried by 10 to 15% of people with northern European Ancestries, with approximately 1 in 150 being p.C282Y homozygous^1^. Iron is an essential element that aids in cell proliferation and growth, but at elevated levels can be toxic. High iron levels have been linked to cancer due to its contribution to tumour initiation and growth^2 3^. The iron overload associated with haemochromatosis is well established in playing a causal role in the development of liver disease and liver cancer^45^. We previously showed in UK Biobank that male p.C282Y homozygotes have a ten-fold increase in the risk of hepatic malignancies compared to those without the mutations^4^. Female p.C282Y homozygotes however have shown no significant increase in risk of hepatic malignancies^4^ perhaps due to partial protection from severe iron overload through menstrual iron losses^6^.

The reports of risk of other types of cancer in individuals with the haemochromatosis-risk mutations is variable, and studies have often not separately examined cancer risk in males and females. A population-based cohort study of almost 7,000 Swedish haemochromatosis patients followed for up to 48 years examined the risk of non-hepatic gastrointestinal cancers compared to the background population. Male and female HH patients had a significantly increased standardized incidence ratio (SIR) of oesophageal squamous cell carcinoma (SIR = 3.2, 95% CI = 1.3–6.6) and colon adenocarcinoma (SIR = 1.4, 95% CI = 1.1–1.9)^7^. However, this study consisted of diagnosed HH patients, with information on *HFE* mutation status unavailable. The Melbourne Collaborative Cohort Study found that p.C282Y homozygotes had an increased risk of colorectal cancer (HR = 2.28; 95% CI = 1.22, 4.25; p = 0.01) and female p.C282Y homozygotes had an increased risk of breast cancer (HR = 2.39; 95% CI = 1.24, 4.61; p = 0.01), but male p.C282Y homozygotes were not at increased risk of prostate cancer^8^. A study of Finnish cancer patients showed that frequency of *HFE* mutations (p.C282Y and p.H63D) did not significantly differ between male prostate cancer patients the population-based controls^9^. A large case-control study in the U.S. did not observe a significant association between haemochromatosis and risk of pancreatic cancer in men and women aged over 66 years^10^.

Given the uncertainty about cancer risks (outside the liver) in community populations with haemochromatosis related genotypes, we estimated the risk of several types of cancer by HH genotype status (p.C282Y/p.H63D) compared to those without the mutations, stratified by sex in a large community genotyped sample of participants of European descent. Clarifying these risks is important given the spread of direct to consumer and clinical testing of asymptomatic individuals for iron related genotypes.

## Methods

Participants were from the UK Biobank cohort, a large prospective study of more than 500,000 community volunteers, who were assessed between 2006 and 2010 at baseline in 22 centers throughout England, Scotland and Wales^11^. Participants were aged 40 to 70 years at baseline and assessment included collecting information on demographics, lifestyle variables and disease history, including providing a blood sample for genotyping. UK Biobank gained ethical approval from the North West Multi-Centre Research Ethics Committee (Research Ethics Committee reference 11/NW/0382). At the baseline assessment, participants gave written informed consent for the collection of data, genotyping from blood samples, and linkage to electronic medical records for follow-up.

### Genotyping

Data were available for 451,143 UK Biobank participants of European descent, with *HFE* p.C282Y (rs1800562) and *HFE* p.H63D (rs1799945) genotype information. UK Biobank used Affymetrix microarrays (800,000 markers directly genotyped); *HFE* p.H63D was directly genotyped on the microarray but *HFE* p.C282Y was imputed by the central UK Biobank team^12^. 98.7% of participants were imputed with 100% confidence. 1.26% were recoded (i.e. estimated genotype dose between 0 and 0.25 set to 0, values between 0.75 and 1.25 set to 1, and finally between 1.75 and 2 to 2) and the remaining 0.04% of participants (n=183) were excluded due to imprecise p.C282Y imputation.

### Cancer Outcomes

Prevalent cancer diagnoses were from baseline self-reported questionnaires, plus hospital inpatient data (National Health Service Hospital Episode Statistics) coded with International Classification of Diseases 10^th^ revision (ICD-10) codes from 1996 to baseline assessment. Incident cancer diagnoses were from baseline assessment (2006 to 2010) to the end of follow-up from hospital inpatient data (to March 2021) and national cancer registry (to July 2019). Information on cancer deaths were obtained from death registrations available until March 2021. We examined any cancer (excluding non-melanoma) and 16 of the most common cancer types: bladder, blood (with sub-analyses of leukemia and lymphoma), bone, brain, breast, colorectal, kidney, lung, melanoma, oesophageal, ovarian, pancreatic, prostate and stomach (see Supplementary Table 1 for ICD-10 coding used). We did not examine liver cancer as we have previously published data on this outcome^4^. Primary care follow-up data were not included for cancer diagnoses as the completeness of case ascertainment in cancer registry data in England is high at around 98-99% ^13^.

### Statistical Analysis

Cox proportional hazards regression models were used to test genotype associations with the risk of incident cancer. Each incident cancer outcome excluded participants with any respective baseline prevalent cancer diagnosis. All models were stratified by sex, and adjusted for age, assessment center, genotyping array and ten genetic principal components generated in European-descent participants, accounting for population genetics sub-structure. We report significant associations based on simple p-values without correction for multiple testing (p<0.05). In addition, we report Bonferroni corrected p-values dividing the 0.05 significance level by 17 cancer types (p<0.003). We estimated lifetable probabilities of incident cancer outcomes from age 40 to 75 years in 5-year bands by sex and *HFE* genotypes, applying observed incidence rates in each age group to a notional cohort, estimating cumulative incidence proportions from age 40 to 75 years. All analyses were performed in Stata version 15.1

## Results

### Characteristics of Participants

We included 451,143 participants of European descent in analyses. The mean age of participants was 56.8 years (standard deviation, SD 8.0, range 40 to 70 years) and 54.3% were female. 2,890 (0.6%) of the cohort were p.C282Y homozygotes. At baseline, 12.1 % of male and 3.4% of female p.C282Y homozygotes were diagnosed with hereditary haemochromatosis. Participants were followed-up from baseline for incident cancer diagnoses for a mean period of 11.6 years (maximum 15.0 years). Non-melanoma skin cancer was not included in our analyses.

In males, 16.9% of p.C282Y homozygotes had an incident diagnosis of any cancer compared to 13.4% in those without mutations. In males, prostate cancer was the most common cancer in both p.C282Y homozygotes (6.8%) and in those with no mutations (5.4%) (Table 1a; Supplementary Table 2a). In females, 9.8% of p.C282Y homozygotes had an incident diagnosis of any cancer compared to 10.2% in those without mutations. In females, breast cancer was the most common cancer in both p.C282Y homozygotes (3.6%) and in those with no mutations (3.9%) (Table 1b; Supplementary Table 2b).

**Table 1.**
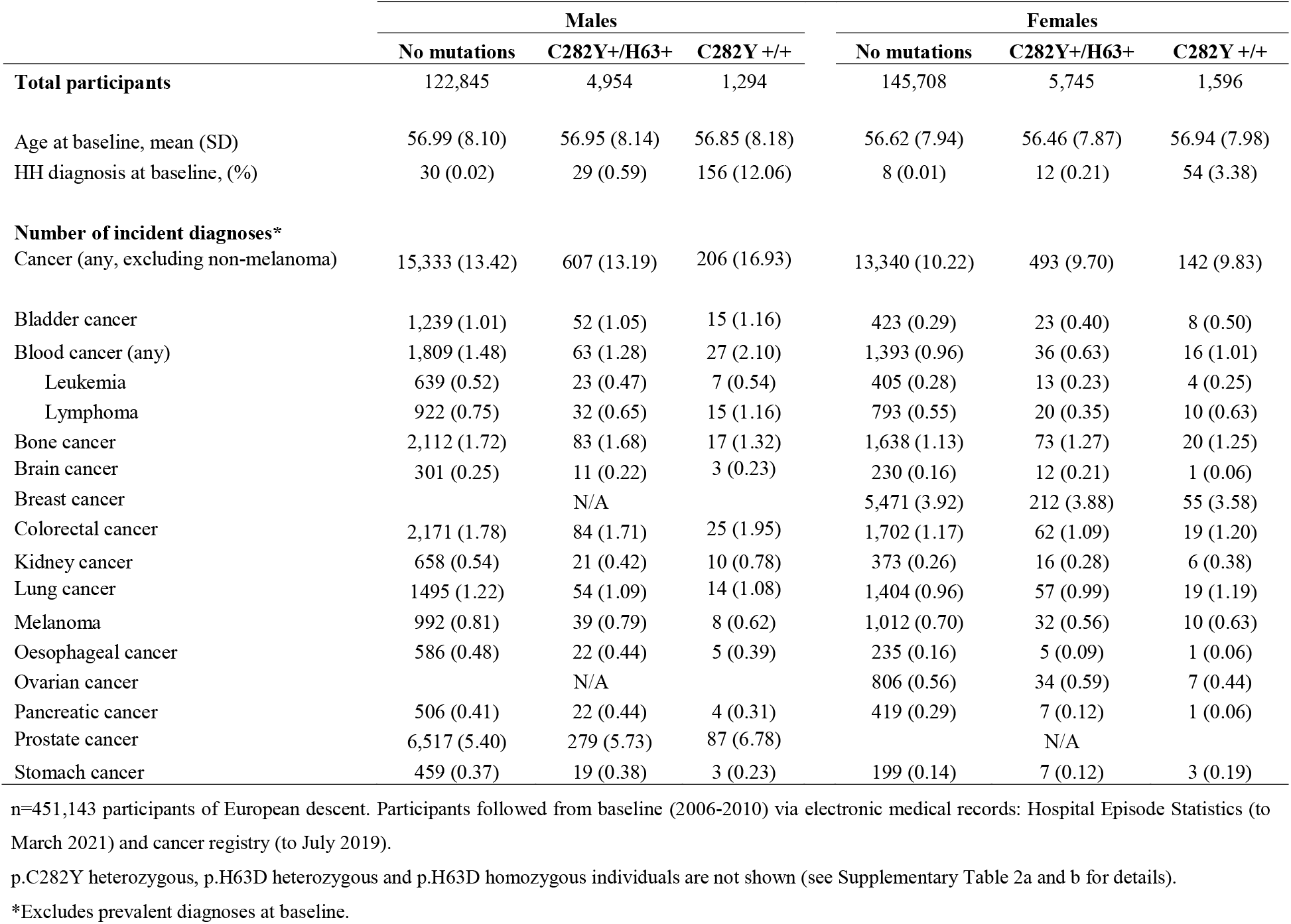
Baseline characteristics of sample and number of incident cancer diagnoses during follow-up, by genotype

### Hazard Ratios for Incident Cancer Outcomes

In male p.C282Y homozygotes, there was a significant increase in the risk of any cancer (HR = 1.29, 95% CI = 1.12-1.48, p=0.0003, n=206). Male p.C282Y homozygotes were also at a significantly increased risk of prostate cancer (HR = 1.32, 95% CI = 1.07-1.63, p = 0.01, n =87) compared to those with no mutations. There is not a significantly increased risk in prostate cancer deaths in male p.C282Y homozygotes, but power was limited due to low case numbers. (HR = 0.73, 95% CI = 0.27-1.94, p = 0.53, n = 4). Male p.C282Y heterozygotes also had a borderline significant increase in risk of prostate cancer (HR = 1.06, 95% CI = 1.00-1.13, p = 0.04, n = 1,381). There was no excess of any type of cancer in male p.C282Y/p.H63D compound heterozygotes. There was also no increased risk of any cancers in female p.C282Y homozygotes or female p.C282Y/p.H63D compound heterozygotes. Unexpectedly, male p.C282Y heterozygotes had a significantly decreased risk of blood cancer (and also the subtypes leukemia and lymphoma). Female p.C282Y/p.H63D compound heterozygotes had a significantly decreased risk of blood cancer and pancreatic cancer. Female p.H63D homozygotes had a significantly lower risk of colorectal cancer. However, following Bonferroni correction for multiple testing, only the association between male p.C282Y homozygotes and an increased risk of any cancer remained significant (see Table 2a and 2b; Supplementary Table 3a and 3b).

**Table 2a.**
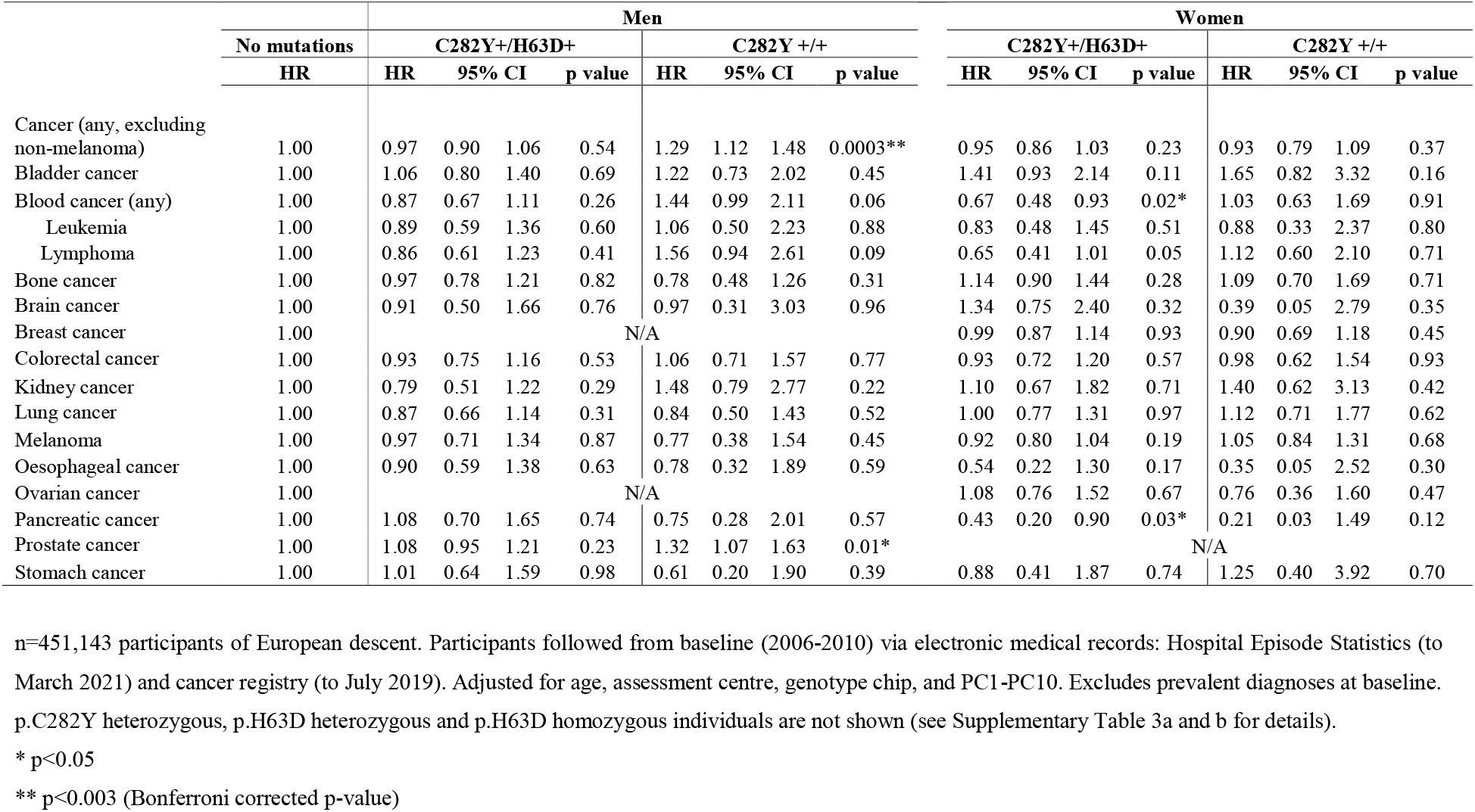
Hazard Ratio (95% CI) for risk of incident cancer by p.C282Y/p.H63D status compared to those with no mutations

### Lifetable Risks of Incident Cancer Diagnoses During Follow-up

In lifetable estimates based on observed incidence rates in 5-year age-band from 40 to 75 years, 14.4% (95% CI = 11.7-17.7) of male p.C282Y homozygotes were projected to develop prostate cancer by age 75, compared to 10.7% (95% CI = 10.4-10.9) in those with no mutations – this is an excess proportion of 3.8% (95% CI = 1.3-6.8).

## Discussion

Our study represents the largest study of community genotyped haemochromatosis participants reported to date, with over 10-fold more p.C282Y homozygotes than previously analyzed^21^. In this UK Biobank study, male *HFE* p.C282Y homozygotes had previously reported markedly increased liver cancer rates^4^ but also a modest 32% increase in the risk of prostate cancer compared to participants without the mutations. In female *HFE* p.C282Y homozygotes, and in male and female p.C282Y/p.H63D compound heterozygotes, there were no significant increases in risk of any types of cancer.

The increased risk of prostate cancer in male p.C282Y homozygotes observed in our study is a novel result which has not been seen previously in other studies. Though the association was not statistically significant after correcting for studying 17 cancer types, Bonferroni correction is known to be conservative and there is considerable *a priori* evidence for *HFE* p.C282Y homozygous males experiencing excess disease. Prostate cancer was the most common cancer in UK Biobank males and is also the most common cancer in the UK for males accounting for more than one quarter of male cases (27%)^17^. We found an excess 3.8% of male p.C282Y homozygotes projected to get prostate cancer by age 75 compared to those without mutations (14.4% compared to 10.7%). In contrast, a study of Finnish cancer patients showed that male p.C282Y homozygotes were not at an increased likelihood of prostate cancer compared to those without the mutations (OR = 1.66, 95% CI = 0.45– 6.16, p = 0.45)^9^. However, this study comprised of a smaller number of 843 p.C282Y homozygotes, of which only 9 had prostate cancer^9^ compared to 87 of 1,294 male p.C282Y homozygotes with prostate cancer in our study. Similarly, the Melbourne Collaborative Cohort Study found no increased risk of prostate cancer in male p.C282Y homozygotes (HR = 0.96; 95% CI 0.43-2.15; p = 0.92)^8^. This study also had a much smaller number of 193 p.C282Y homozygotes (of which only 6 had prostate cancer) but were a very similar age with 99.3% also being aged 40 to 69 years^8^.

Beyond the previously reported increase in liver cancer^4^ and the increase in prostate cancer seen here, we found no significant increase in risk of any other type of cancer in male or female p.C282Y homozygotes. We found no association between HFE-genotypes and the risk of pancreatic cancer, despite the previous suggestion that higher serum iron is associated with pancreatic cancer^18^. In contrast to our results, a case control study in a Chinese Han population found that with each additional copy of p.H63D allele there was a 21% increase in risk of pancreatic cancer (OR□=□1.21, 95 % CI 1.05–1.39, *P*□=□7.72□×□10^−3^) ^19^. However, in line with our results a large case-control study in the U.S. also did not find a significant association between haemochromatosis and risk of pancreatic cancer, in older men and women^10^. We also found no association between p.C282Y homozygous status and colorectal cancer, although the Melbourne Collaborative Cohort Study found over a two-fold increase in risk. However, the Melbourne study included just 193 p.C282Y homozygotes compared to 2,890 in the current study. ^8^ In male p.C282Y heterozygotes we found a significant but modest decrease in the risk of blood cancers which has not been seen before, and therefore warrants further investigation. In women, we found no association between *HFE*-genotypes and the risk of breast cancer in our study despite a previous study in Sweden showing a significant positive association between serum iron and breast cancer risk in postmenopausal women^20^, and an Australian cohort study which showed a 2.4-fold increase in the risk of breast cancer in female p.C282Y homozygotes^8^.

## Strengths and Limitations

This was a large study of community genotyped participants, with more than ten times the number of p.C282Y homozygotes compared to the previous largest study^21^. The study had objective classification of cancer diagnoses from hospital inpatient data, and also the cancer registry which has high levels of case ascertainment completeness (98-99%) ^13^. There was a long prospective follow-up period of over a mean of 11 years, in 40 to70 year olds. However, the study has some limitations. Firstly, at baseline UK Biobank participants were healthier than the general population^22^; however risk estimates are from incident cancer during follow-up so this should be less susceptible to any baseline bias. Secondly, we included participant of European ancestry so results may not be applicable to other populations. Third data on ferritin concentrations and transferrin saturation were not available in UK Biobank so we do not have information to comment on iron loading specifically. Fourth, although our findings were adjusted for chronological age, we did not evaluate heterogeneity between individuals in terms of extent or rate of biological aging which may interact with alterations in iron-mediated cellular metabolism associated with specific HH genotypes in determining cancer risk.

## Conclusions

In male p.C282Y homozygotes, beyond the known increased in risk of liver cancer, there was also a modest increase in the risk of prostate cancer. Female p.C282Y homozygotes, and male and female p.C282Y/p.H63D compound heterozygotes were not at an increased risk of any types of cancer.

## Data Availability

Data are available on application to the UK Biobank (https://www.ukbiobank.ac.uk/enable-your-research/register).

## Abbreviations

HR: hazard ratio
HH: hereditary haemochromatosis
ICD-10: International Classification of Diseases 10^th^ revision
OR: odds ratio
SD: standard deviation
SIR: standardized incident ratio;

## Conflict of interest disclosures

None reported.

## Acknowledgements

This research was conducted using the UK Biobank resource, under application 14631. We thank the UK Biobank participants and coordinators for the dataset.

## Funding

This research was funded by an award to DM by the UK Medical Research Council (MR/S009892/1). JA is supported by an NIHR Advanced Fellowship (NIHR301844). DM and LCP are supported by the University of Exeter Medical School. SVT and FMT acknowledge support from the National Cancer Institute and (R01 CA188025 [SVT] and R01 CA233636 [FMT]). GAK acknowledges support from the National Institute on Aging (P30 AG067988; R33 AG061456), as well as the Travelers Chair in Geriatrics and Gerontology.

## Role of the Funder

The funders had no role in the design and conduct of the study; collection, management, analysis, and interpretation of the data; preparation, review, or approval of the manuscript; and decision to submit the manuscript for publication.

---

**Figure 1.**
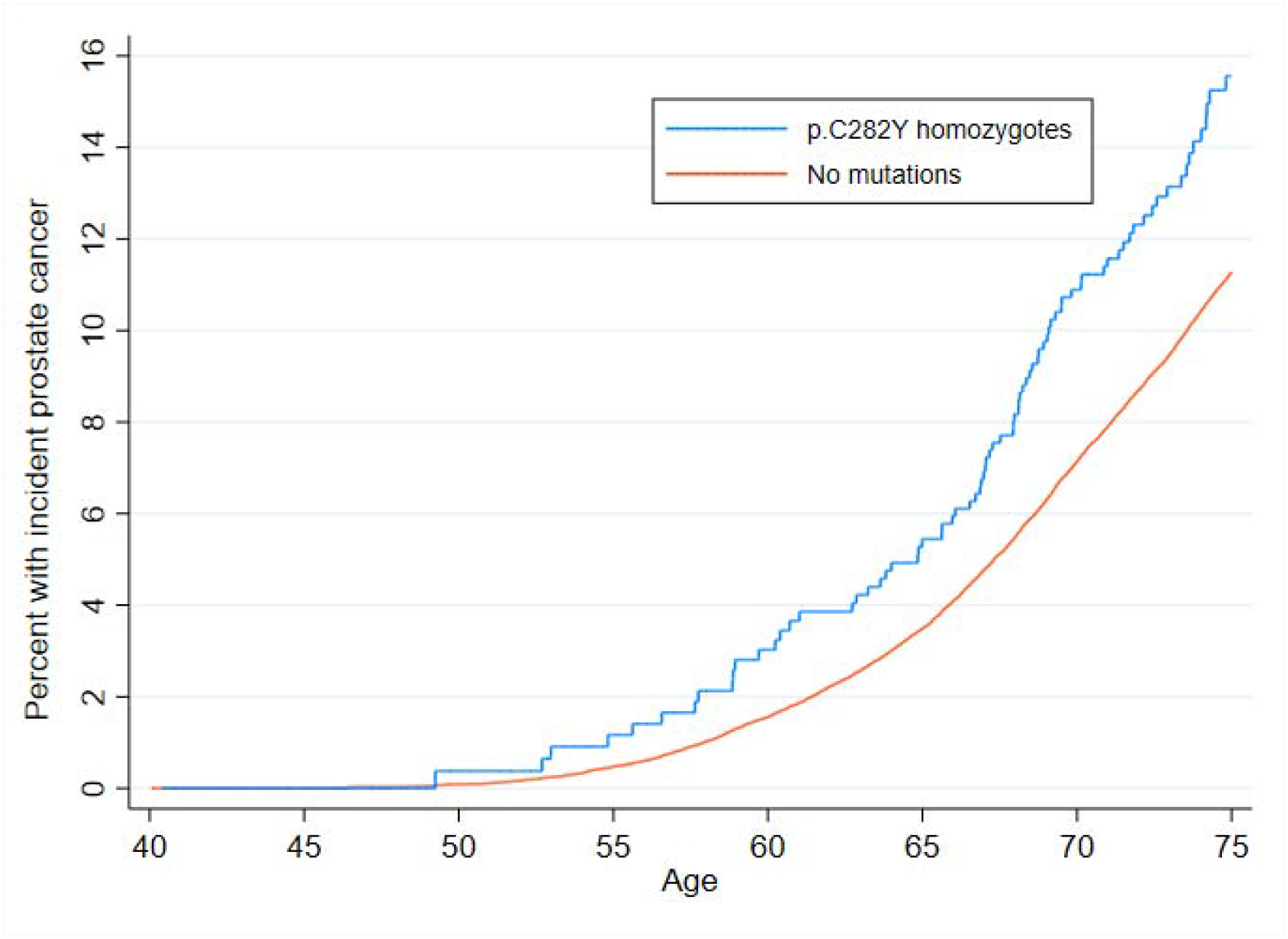
Kaplan-Meier curve for the incidence of prostate cancer in male *HFE* p.C282Y homozygotes compared to those with no mutations Participants followed from baseline (2006-2010) via electronic medical records: Hospital Episode Statistics (to March 2021) and cancer registry (to July 2019). Excludes prevalent diagnoses at baseline.

**Supplementary Table 1.**
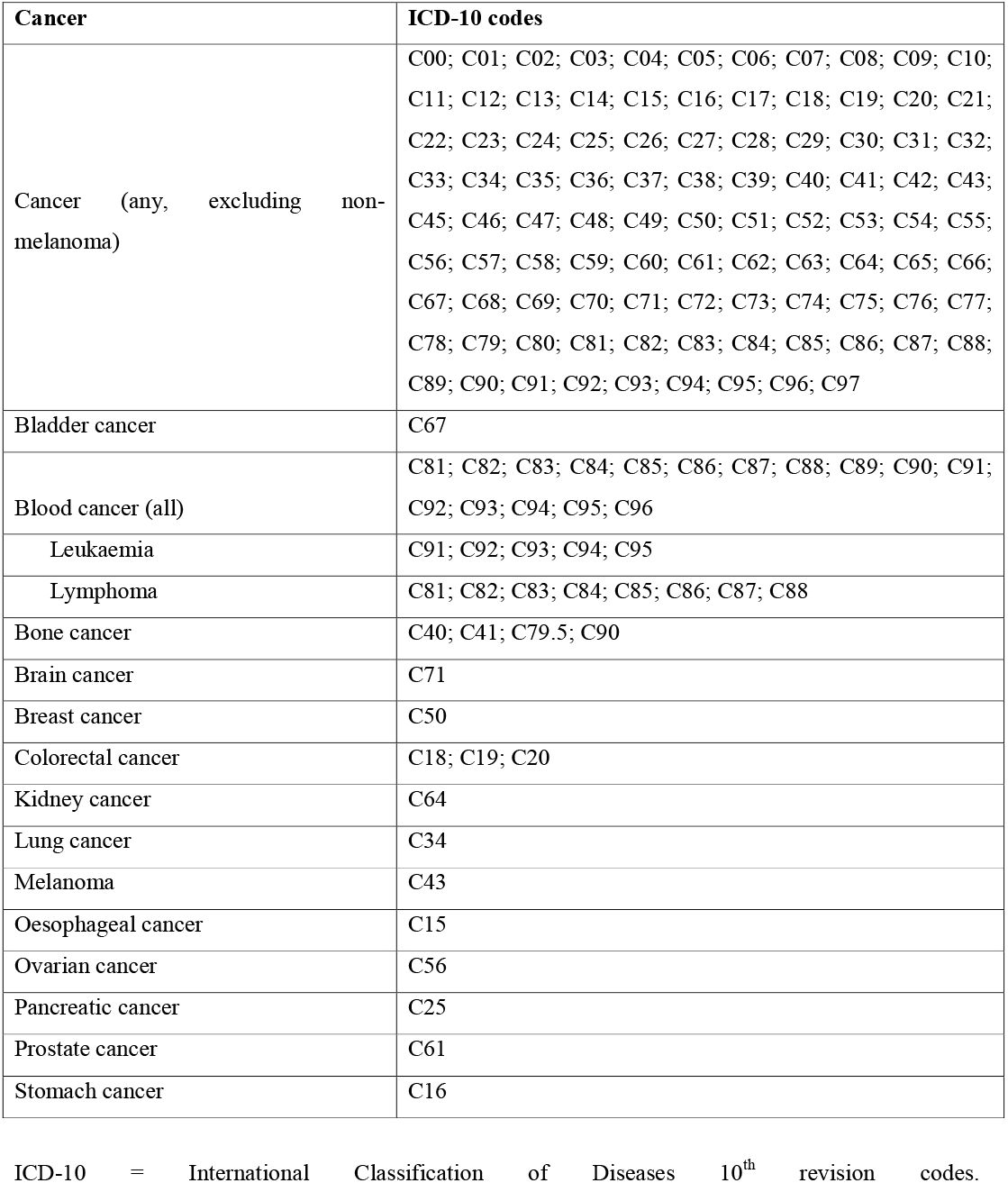
Cancer disease coding

**Supplementary Table 2a.** Baseline characteristics of sample and number of incident cancer diagnoses during follow-up in males, by genotype

|  | Males |  |  |  |  |  | Total |
| --- | --- | --- | --- | --- | --- | --- | --- |
|  | No mutations | H63D+/- | H63D+/+ | C282Y+/H63+ | C282Y+/- | C282Y +/+ |  |
| **Total participants** | 122,845 | 47,988 | 4,674 | 4,954 | 24,575 | 1,294 | 206,330 |
| Age at baseline, mean (SD) | 56.99 (8.10) | 57.02 (8.13) | 56.99 (8.08) | 56.95 (8.14) | 57.02 (8.12) | 56.85 (8.18) | 57.0 (8.11) |
| HH diagnosis at baseline, (%) | 30 (0.02) | 17 (0.04) | 8 (0.17) | 29 (0.59) | 27 (0.11) | 156 (12.06) | 267 (0.13) |
| **Number of incident diagnoses\*** |  |  |  |  |  |  |  |
| Cancer (any, excluding non-melanoma) | 15,333 (13.42) | 6,164 (13.78) | 566 (13.07) | 607 (13.19) | 3,143 (13.72) | 206 (16.93) | 26,019 (13.55) |
| Bladder cancer | 1,239 (1.01) | 501 (1.05) | 56 (1.21) | 52 (1.05) | 270 (1.10) | 15 (1.16) | 2,133 (1.04) |
| Blood cancer (any) | 1,809 (1.48) | 683 (1.43) | 62 (1.33) | 63 (1.28) | 300 (1.23) | 27 (2.10) | 2,944 (1.44) |
| Leukemia | 639 (0.52) | 273 (0.57) | 17 (0.36) | 23 (0.47) | 103 (0.42) | 7 (0.54) | 1,062 (0.52) |
| Lymphoma | 922 (0.75) | 337 (0.70) | 33 (0.71) | 32 (0.65) | 148 (0.60) | 15 (1.16) | 1,487 (0.72) |
| Bone cancer | 2,112 (1.72) | 822 (1.72) | 85 (1.82) | 83 (1.68) | 416 (1.70) | 17 (1.32) | 3,535 (1.72) |
| Brain cancer | 301 (0.25) | 129 (0.27) | 7 (0.15) | 11 (0.22) | 69 (0.28) | 3 (0.23) | 520 (0.25) |
| Breast cancer |  |  | N/A |  |  |  |  |
| Colorectal cancer | 2,171 (1.78) | 873 (1.83) | 71 (1.53) | 84 (1.71) | 434 (1.78) | 25 (1.95) | 3,658 (1.79) |
| Kidney cancer | 658 (0.54) | 264 (0.55) | 23 (0.49) | 21 (0.42) | 150 (0.61) | 10 (0.78) | 1,126 (0.55) |
| Lung cancer | 1,495 (1.22) | 608 (1.27) | 46 (0.99) | 54 (1.09) | 302 (1.23) | 14 (1.08) | 2,519 (1.22) |
| Melanoma | 992 (0.81) | 398 (0.84) | 41 (0.88) | 39 (0.79) | 213 (0.87) | 8 (0.62) | 1,691 (0.83) |
| Oesophageal cancer | 586 (0.48) | 248 (0.52) | 30 (0.64) | 22 (0.44) | 135 (0.55) | 5 (0.39) | 1,026 (0.50) |
| Ovarian cancer |  |  | N/A |  |  |  |  |
| Pancreatic cancer | 506 (0.41) | 193 (0.40) | 18 (0.39) | 22 (0.44) | 107 (0.44) | 4 (0.31) | 850 (0.41) |
| Prostate cancer | 6,517 (5.40) | 2,608 (5.52) | 239 (5.20) | 279 (5.73) | 1,381 (5.71) | 87 (6.78) | 11,111 (5.47) |
| Stomach cancer | 459 (0.37) | 169 (0.35) | 19 (0.41) | 19 (0.38) | 98 (0.40) | 3 (0.23) | 767 (0.37) |

**Supplementary Table 2b.** Baseline characteristics of sample and number of incident cancer diagnoses during follow-up in females, by genotype

|  | Females |  |  |  |  |  | Total |
| --- | --- | --- | --- | --- | --- | --- | --- |
|  | No mutations | H63D+/- | H63D+/+ | C282Y+/H63+ | C282Y+/- | C282Y +/+ |  |
| **Total participants** | 145,708 | 57,023 | 5,584 | 5,745 | 29,157 | 1,596 | 244,813 |
| Age at baseline, mean (SD) | 56.62 (7.94) | 56.58 (7.93) | 56.59 (8.08) | 56.46 (7.87) | 56.49 (7.98) | 56.94 (7.98) | 56.60 (7.94) |
| HH diagnosis at baseline, (%) | 8 (0.01) | 6 (0.01) | 2 (0.04) | 12 (0.21) | 5 (0.02) | 54 (3.38) | 87 (0.04) |
| **Number of incident diagnoses\*** |  |  |  |  |  |  |  |
| Cancer (any, excluding non-melanoma) | 13,340 (10.22) | 5,261 (10.30) | 502 (10.06) | 493 (9.70) | 2,648 (10.15) | 142 (9.83) | 22,386 (10.21) |
| Bladder cancer | 423 (0.29) | 148 (0.26) | 16 (0.29) | 23 (0.40) | 78 (0.27) | 8 (0.50) | 696 (0.28) |
| Blood cancer (any) | 1,393 (0.96) | 531 (0.94) | 57 (1.02) | 36 (0.63) | 283 (0.97) | 16 (1.01) | 2,316 (0.95) |
| Leukemia | 405 (0.28) | 172 (0.30) | 22 (0.39) | 13 (0.23) | 93 (0.32) | 4 (0.25) | 709 (0.29) |
| Lymphoma | 793 (0.55) | 286 (0.50) | 25 (0.45) | 20 (0.35) | 150 (0.52) | 10 (0.63) | 1,284 (0.53) |
| Bone cancer | 1,638 (1.13) | 593 (1.04) | 48 (0.86) | 73 (1.27) | 325 (1.12) | 20 (1.25) | 2,697 (1.10) |
| Brain cancer | 230 (0.16) | 94 (0.16) | 8 (0.14) | 12 (0.21) | 40 (0.14) | 1 (0.06) | 385 (0.16) |
| Breast cancer | 5,471 (3.92) | 2,214 (4.05) | 228 (4.27) | 212 (3.88) | 1,058 (3.79) | 55 (3.58) | 9,238 (3.94) |
| Colorectal cancer | 1,702 (1.17) | 686 (1.21) | 46 (0.83) | 62 (1.09) | 353 (1.22) | 19 (1.20) | 2,868 (1.18) |
| Kidney cancer | 373 (0.26) | 152 (0.27) | 15 (0.27) | 16 (0.28) | 89 (0.31) | 6 (0.38) | 651 (0.27) |
| Lung cancer | 1,404 (0.96) | 560 (0.98) | 57 (1.02) | 57 (0.99) | 286 (0.98) | 19 (1.19) | 2,383 (0.97) |
| Melanoma | 1,012 (0.70) | 388 (0.69) | 34 (0.61) | 32 (0.56) | 195 (0.67) | 10 (0.63) | 1,671 (0.69) |
| Oesophageal cancer | 235 (0.16) | 99 (0.17) | 8 (0.14) | 5 (0.09) | 46 (0.16) | 1 (0.06) | 394 (0.16) |
| Ovarian cancer | 806 (0.56) | 285 (0.50) | 27 (0.49) | 34 (0.59) | 149 (0.51) | 7 (0.44) | 1,308 (0.54) |
| Pancreatic cancer | 419 (0.29) | 164 (0.29) | 16 (0.29) | 7 (0.12) | 104 (0.36) | 1 (0.06) | 711 (0.29) |
| Prostate cancer |  |  |  | N/A |  |  |  |
| Stomach cancer | 199 (0.14) | 73 (0.13) | 5 (0.09) | 7 (0.12) | 43 (0.15) | 3 (0.19) | 330 (0.13) |

n=451,143 participants of European descent. Participants followed from baseline (2006-2010) via electronic medical records: Hospital Episode Statistics (to March 2021) and cancer registry (to July 2019).
\*Excludes prevalent diagnoses at baseline.

**Supplementary Table 3a.**
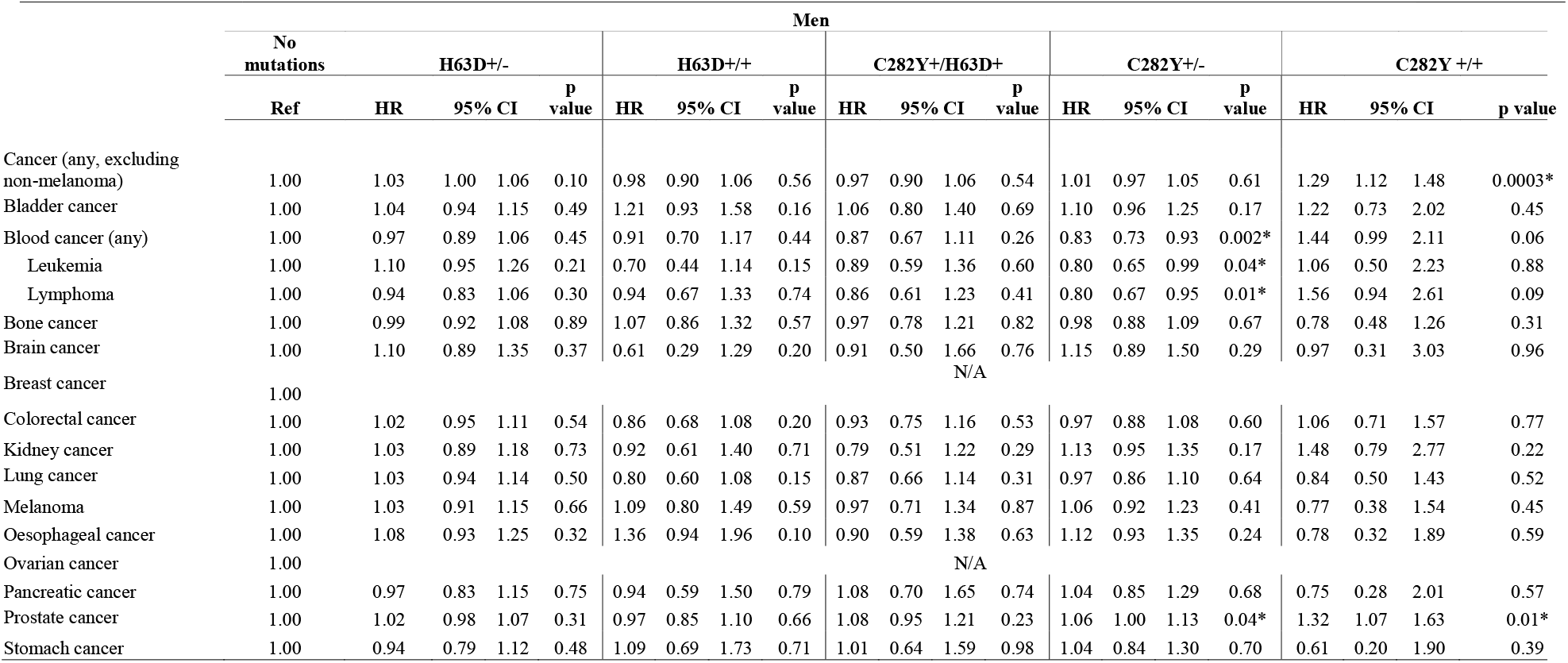
Hazard Ratio (95% CI) for risk of incident cancer by p.C282Y/p.H63D status compared to those with no mutations in males

**Supplementary Table 3b.**
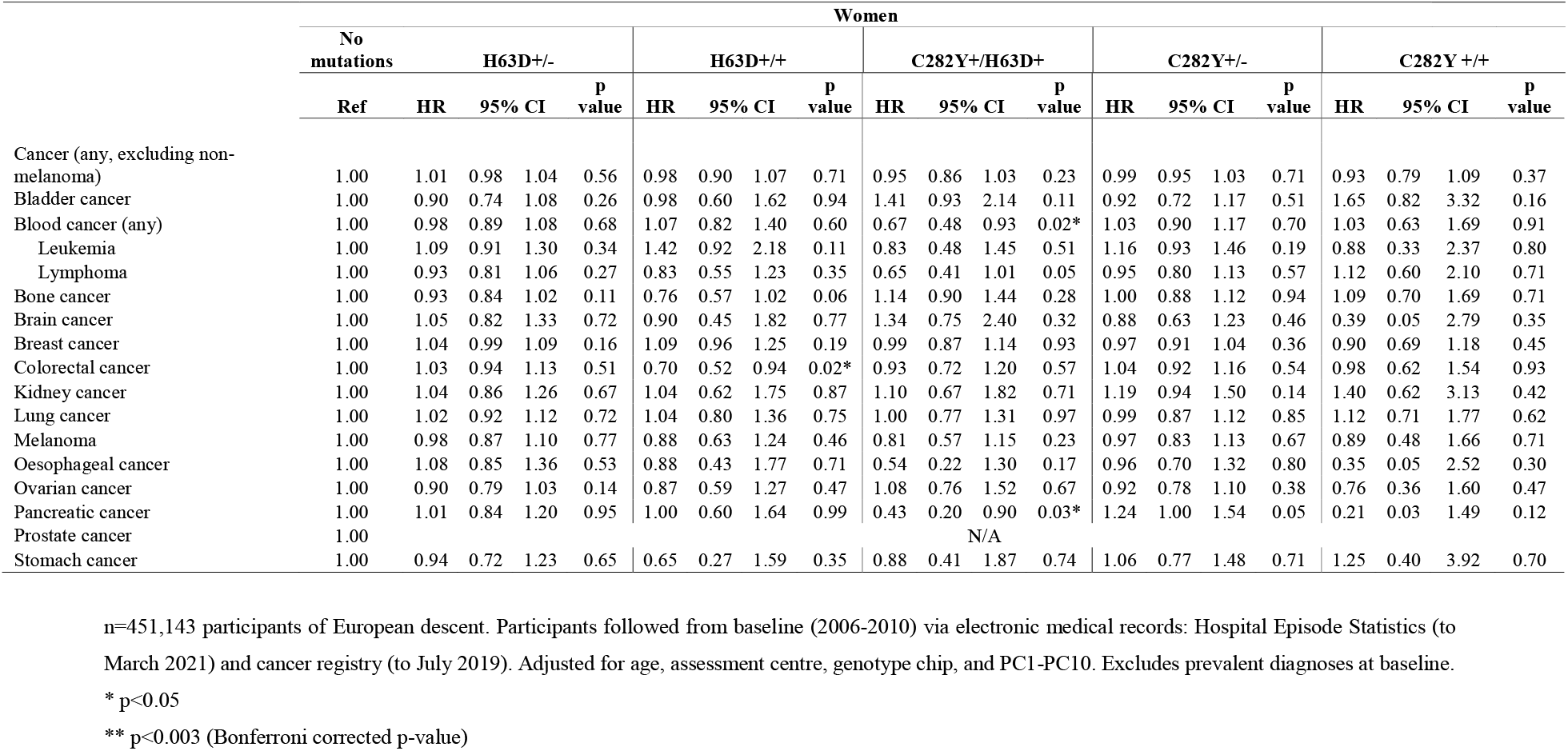
Hazard Ratio (95% CI) for risk of incident cancer by p.C282Y/p.H63D status compared to those with no mutations in females

## Notes

### Competing Interest Statement

The authors have declared no competing interest.

